# The emergence, spread and vanishing of a French SARS-CoV-2 variant exemplifies the fate of RNA virus epidemics and obeys the Black Queen rule

**DOI:** 10.1101/2022.01.04.22268715

**Authors:** Philippe Colson, Philippe Gautret, Jeremy Delerce, Hervé Chaudet, Pierre Pontarotti, Patrick Forterre, Raphael Tola, Marielle Bedotto, Léa Delorme, Anthony Levasseur, Jean-Christophe Lagier, Matthieu Million, Nouara Yahi, Jacques Fantini, Bernard La Scola, Pierre-Edouard Fournier, Didier Raoult

## Abstract

The nature and dynamics of mutations associated with the emergence, spread and vanishing of SARS-CoV-2 variants causing successive waves are complex^1-5^. We determined the kinetics of the most common French variant (“Marseille-4”) for 10 months since its onset in July 2020^5^. Here, we analysed and classified into subvariants and lineages 7,453 genomes obtained by next-generation sequencing. We identified two subvariants, Marseille-4A, which contains 22 different lineages of at least 50 genomes, and Marseille-4B. Their average lifetime was 4.1±1.4 months, during which 4.1±2.6 mutations accumulated. Growth rate was 0.079±0.045, varying from 0.010 to 0.173. All the lineages exhibited a “gamma” distribution. Several beneficial mutations at unpredicted sites initiated a new outbreak, while the accumulation of other mutations resulted in more viral heterogenicity, increased diversity and vanishing of the lineages. Marseille-4B emerged when the other Marseille-4 lineages vanished. Its ORF8 gene was knocked out by a stop codon, as reported in several mink lineages and in the alpha variant. This subvariant was associated with increased hospitalization and death rates, suggesting that ORF8 is a nonvirulence gene. We speculate that the observed heterogenicity of a lineage may predict the end of the outbreak.

## Introduction

The shape of epidemic curves of acute infectious diseases is the subject of several hypotheses and interpretations. The occurrence of successive waves of SARS-CoV-2 infections during the current pandemic was linked to the emergence of viral variants^1-5^, while possible causes of the extinction of epidemics are viral load decrease^6^, herd immunity^7^ (as hypothesized for influenza viruses)^8^, or the implementation of treatment or vaccination^9^. However, the factors and mechanisms involved in the rise and fall of SARS-CoV-2 variants have not been elucidated. We identified in July 2020 at IHU Méditerranée Infection, Marseille, France (which generated 20% of the genomes deposited in GISAID (https://www.gisaid.org/)^10^ by France as on 16/12/2021), a new SARS-CoV-2 variant, named Marseille-4 (later classified as lineage 20A.EU2 and B.1.160 in Nextstrain and Pangolin classifications)^11^. This variant is characterized by 20 mutations, including 13 specific compared with the Wuhan-Hu-1 isolate^5,11^. Seven mutations are nonsynonymous, including one in the spike glycoprotein (S477N). We analysed the epidemiological source and features of this variant and accumulated genetic data through extensive SARS-CoV-2 genomic surveillance by next-generation sequencing from its onset until its disappearance 10 months later in April of 2021. Thus, we could study the nature and dynamics of mutations associated with its emergence, spread, and vanishing.

## Results

### Kinetics of the Marseille-4 variant infection

The identification of the Marseille-4 variant in late July 2020^11^ (Figure 1a) led to design a specific qPCR to evaluate its incidence and 9,616 cases were identified. It was the third most commonly diagnosed variant at our institution after variants Alpha/B.1.160 (n= 10,139) and Delta/B.1.617.2 (11,060). By contrast, it was rarely observed during this period in the UK or Spain, where the Marseille-2/B.1.177 variant predominated (Figure 1b). The shape of the incidence curve of Marseille-4 differed from that of mutants of the Wuhan-Hu-1 isolate or of alpha and delta variants that show a “gamma” distribution^12^. The curve of Marseille-4 included several peaks, with a first in September (week 37), a second in October (week 43), and a third in January 2021 (week 2) before the variant vanished in April 2021 (Figure 1a).

**Figure 1.**
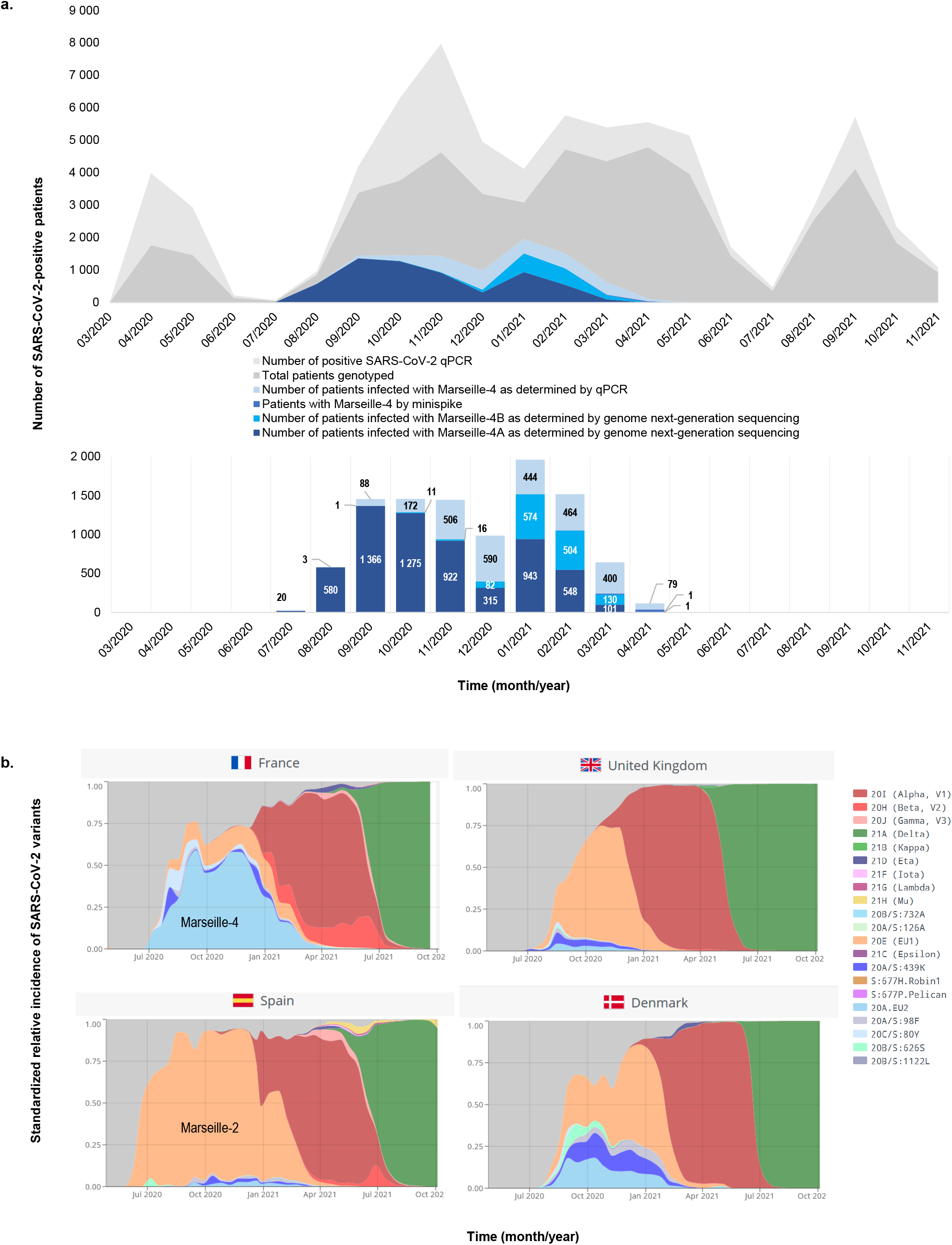
Weekly incidence of SARS-CoV-2 diagnoses at the IHU Méditerranée Infection, Marseille, France and incidence of the SARS-CoV-2 Marseille-4 variant (a) and spread of the Marseille-4 variant in France and three additional European countries (b). Figure 1b is adapted from screenshots from the CoVariants website (https://covariants.org/)^31^.

### Marseille-4 subvariants and lineages

Next-generation sequencing was performed when cycle threshold values (Ct) of qPCR was <30 and a genome was obtained from 7,453 patients (Figure 1a). Phylogenetic analysis and comparative genomics identified two subvariants, i.e. new variants issued from a circulating variant (Marseille-4A and Marseille-4B). The Marseille-4A subvariant contained 22 different lineages of at least 50 SARS-CoV-2 genomes harbouring one to four hallmark nucleotide changes (Figure 2a; Supplementary Tables S1, S2, S3). Interestingly, the single sequence reported from an infected mink farm in France was a Marseille-4A variant^11^. The growth rate varied throughout the Marseille-4 epidemic for each subvariant and lineage (Figures 2b-g; Supplementary Figure S1; Supplementary Tables S2, S3). It was 0.079±0.045 on average and varied from 0.010 for the Marseille-4A.17 lineage to 0.173 for the Marseille-4A.15 lineage. Thus, we observed heterogeneous growth rates for Marseille-4 subvariants and lineages, as indicated by a very high ratio of true heterogeneity to the total observed variation (I^2^ = 99%; p < 0.05; Supplementary Figure S2).

**Figure 2.**
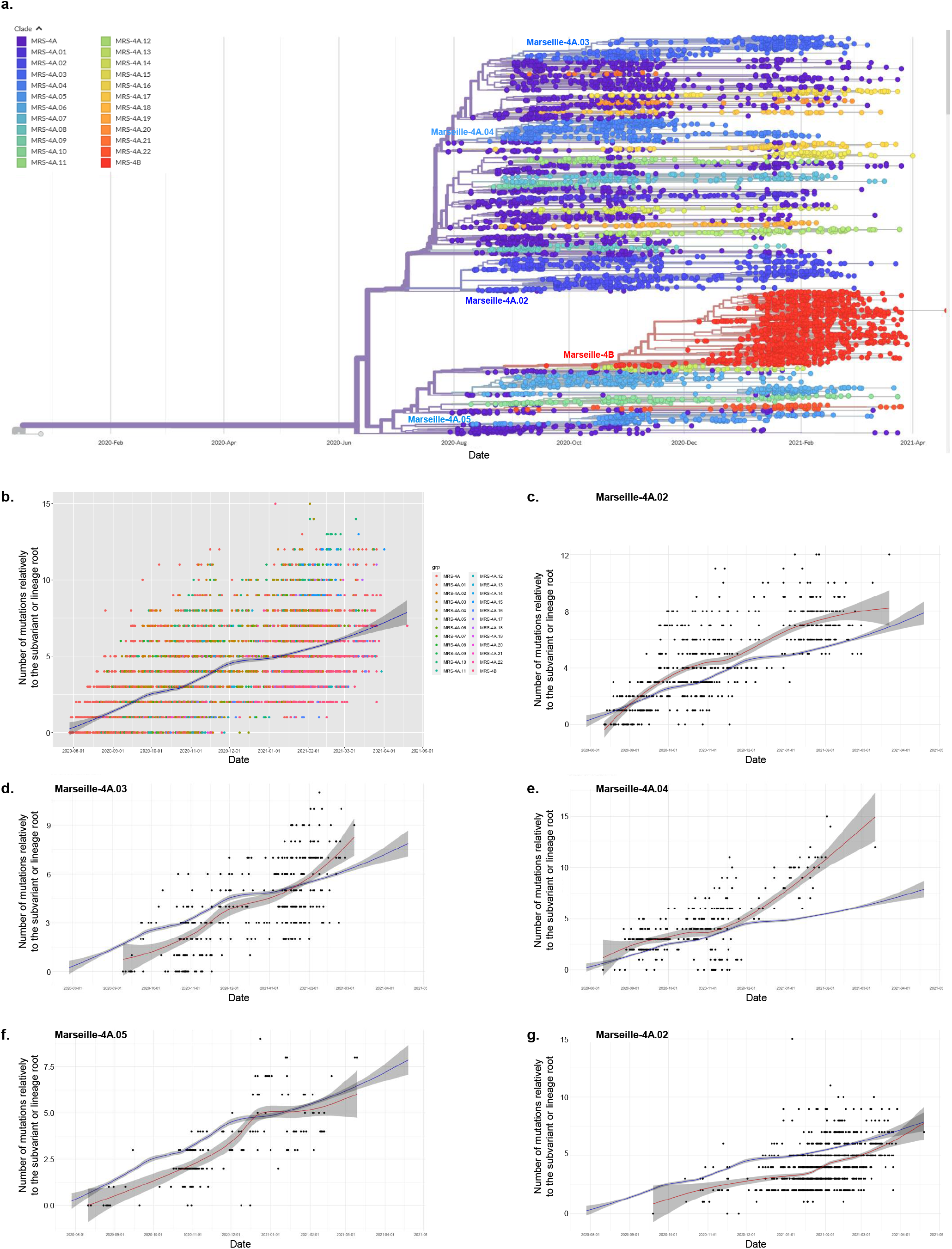
Phylogenetic tree of SARS-CoV-2 Marseille-4 genome sequences obtained from patients diagnosed with SARS-CoV-2 infection at IHU Méditerranée Infection (a) and growth rate of the Marseille-4 variant overall (b) and of four Marseille-4A lineages (c-f) and of the Marseille-4B lineage (g). b-g: Time series of the number of mutations from the variant root of each subvariant or lineage along with the loess (locally estimated scatterplot smoothing) regression curve and its 95% confidence interval.

### ORF8 gene inactivation in the Marseille-4B subvariant

The Marseille-4B subvariant spread from September 2020 to March 2021 with a gamma distribution of cases (Figure 1a; Supplementary Tables S2, S3). It expanded significantly in December 2020 during the vanishing of the other lineages. It exhibited a mean number of 4.1±1.6 lineage-specific mutations (range, 0-15; n= 1,319 genomes). Interestingly, the ORF8 gene was knocked out at the origin of the Marseille-4B variant. This gene may play a key role in immune modulation and increases virus multiplication^13^. Its inactivation by a stop codon has been reported in several mink lineages and in all genomes of the Alpha variant. The dimeric structure of the ORF8 protein (wild-type and mutant forms) is shown in Figure 3a-c. The dimer is stabilized by a covalent bond (a disulfide bridge) between two cysteine residues in the N-terminal region of each subunit^14^. Mutation A65S does not induce major structural or electrostatic surface potential alterations (Figure 3b). Both the initial A65 and mutant A65S residues are well exposed to the solvent and occupy approximately the same volume. By contrast, the truncated 18-63 form leads to a different protein, despite its sequence identity with the 18-63 region of the initial ORF8 protein chains (Figure 3c). The electrostatic surface potential of the truncated protein is also significantly affected, with an increase in both neutral and electronegative surface areas. These structural data suggest a total loss of ORF8 function for the truncated 18-63 form. Finally, mutation H17Y affects the C-terminal residue of the signal peptide, so it is not present in the mature form of ORF8, as shown in Figure 3a-c.

**Figure 3.**
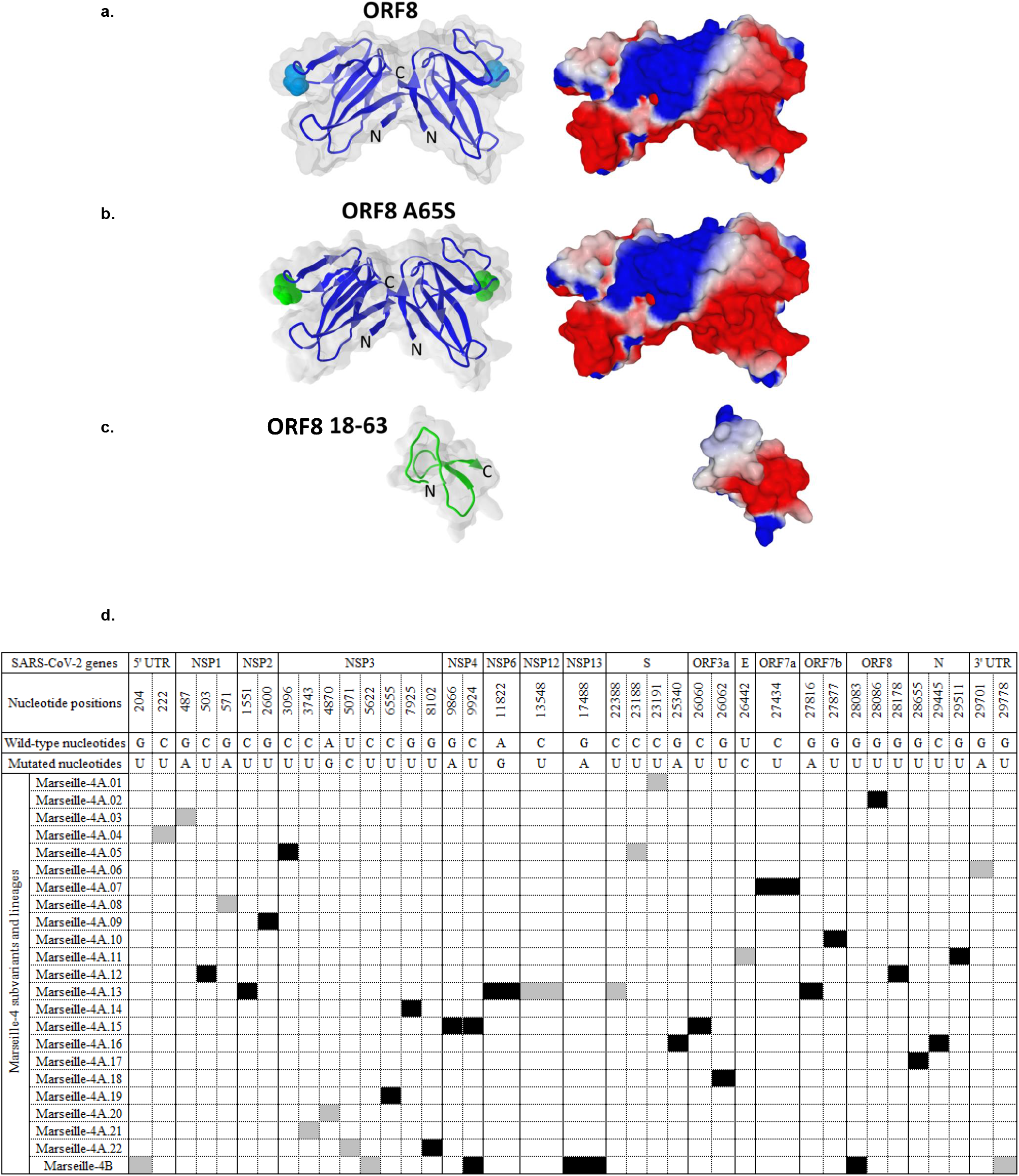
Structure of the SARS-CoV-2 ORF8 protein and its mutated and truncated forms (a-c), and signature mutations in the genomes obtained for each of the Marseille-4 subvariants and lineages (d). a-c: The upper panel (a) shows the structure of dimeric SARS-CoV-2 ORF8 shown as superimposed surface and cartoon representations. The missing amino acids (65-66 in chain A and 66-68 in chain B) were inserted with Swiss-PdbViewer in PDB: 7JTL, and the resulting model was minimized using the Polak-Ribiere algorithm of Hyperchem. Residue A65 in both chains is highlighted in cyan. The structure of the ORF8 mutant A65S (highlighted in green) was modelled using Swiss-PdbViewer and Hyperchem (middle panels) (b). The structure of truncated ORF8 18-63 (bottom panels) was obtained using Hyperchem (c). For all models, the surface potential of the protein is shown in the right panels (blue, positive; red, negative; white, neutral). b: Synonymous nucleotide changes are indicated by a grey background. Non-synonymous nucleotide changes are indicated by a black background.

### Severity of the Marseille-4B subvariant infections

We compared the characteristics of the first 181 patients identified as infected with Marseille-4B and 1,647 patients identified as infected with Marseille-4A (Table 1a-b). Patients infected with Marseille-4B were more likely to be female and older than those infected with Marseille-4A. The mean Ct values did not differ between the two groups of patients. Higher hospitalization and death rates were observed in patients infected with Marseille-4B. Multivariate analysis (Table 1b) confirmed that increased hospitalization rate was significantly associated with male sex, older age, a lower viral load (increased Ct value) and Marseille-4B infection. An increased rate of transfer to the intensive care unit was significantly associated with male sex and older age. An increased death rate was significantly associated with male sex, older age and Marseille-4B infection. Thus, we concluded that Marseille-4B was more virulent and suspected that it is related to the knock-out of ORF8.

**Table 1.**
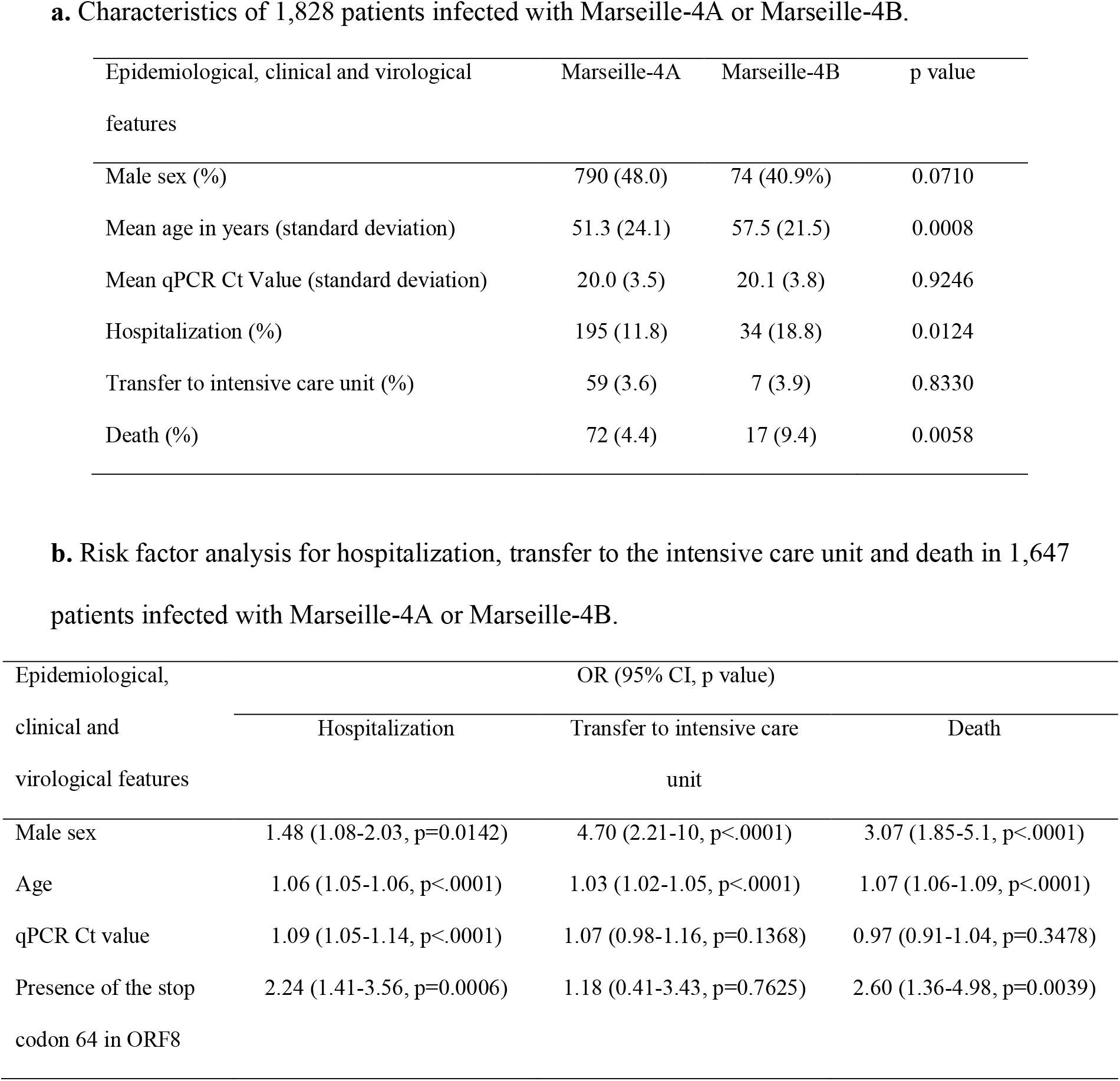
Characteristics of 1,828 patients infected with Marseille-4A or Marseille-4B (a) and risk factor analysis for hospitalization, transfer to the intensive care unit and death in 1647 patients infected with Marseille-4A or Marseille-4B (b).

### Some mutations are associated with expansion of Marseille-4A lineages, other with their vanishing

Analysing the kinetics of new variants without enough viral sequences leads to confused interpretation because of the superposition of different lineages in this clade. As for Marseille-4, we identified nucleotide changes among the 22 Marseille-4A lineages. Most of the Marseille-4 subvariants and lineages showed a gamma distribution of cases with a lifetime of 4.1±1.4 months on average (Supplementary Figure S3), during which 4.1±2.6 mutations accumulated in viral genomes. RNA viruses have a high mutation rate^15,16^ and SARS-CoV-2 accumulates approximately one mutation every 2 weeks^17^. We found signature mutations in each of these lineages, including in the Nsp1, Nsp2, NSp2, spike, ORF7a, ORF7b, and ORF8 genes (Supplementary Table S1; Figure 3d). Interestingly, the accumulation of mutations resulting in an increasing genetic divergence correlated with a decreased incidence. Thus, the accumulation of nonlethal and nonfavouring mutations leads gradually to a dispersion of the lineages, a decrease in viral fitness and vanishing of the epidemic.

## DISCUSSION

Here, we described the complete cycle of emergence, spread and vanishing of the Marseille-4 variant identified in France from mink^11^ by analysing 7,453 genomes. The viral mutation rate leads to the accumulation of many random mutations, most presumably mildly deleterious, with little effect on fitness. Only 10^−8^ may be associated with a fitness gain^15,18,19^. Overall, the RNA virus fitness evolution includes an initial period of rapid multiplication possibly caused by a positive mutation followed by the decline of viral fitness caused by accumulation of unfit mutations, as described for the vesicular stomatitis virus^15,20^.

We observed heterogeneity of the growth rates for the different Marseille-4 subvariants and lineages, making it challenging to generalize the behaviour of one SARS-CoV-2 subvariant or lineage to all of them. All Marseille-4 lineages present a genetic signature with mutations sometimes associated with the inactivation of ORF7a or ORF7b, as described^21^. None of these mutations, apart from those located in the spike gene, were predicted to be possibly associated with increased transmissibility. Knock-out of the ORF8 and ORF7a/b genes shows that SARS-CoV-2 virulence may be associated with a loss of genes, as shown for bacteria in which the decline in genomic content is often associated with an increased specificity and virulence^22^. Analysis of the Marseille-4B subvariant revealed the presence of a stop codon in ORF8, which was observed in the alpha variant and viruses infecting minks and pangolins^23^. Marseille-4B subvariant is more virulent, at a time when the other Marseille-4 lines had vanished or were vanishing. This finding suggests that this gene may be a so-called “nonvirulence gene”^24^ and that SARS-CoV-2 may follow the “Black Queen” hypothesis, which proposes that the loss of a gene may provide a selective advantage when its function is dispensable^25^, leading to the uselessness of correcting inactivating mutations on the “Use it or lose it” model^26^. As expected, structural analysis of the truncated form of ORF8 (ORF8 18-63) confirms that the large deletion induced by the stop codon results in a distinct protein that does not retain any resemblance to the native ORF8 dimer. Interestingly, partial or complete deletions of the ORF8 gene were observed during the middle and late phases of the SARS-CoV epidemic in 2002-2003^27^. Additionally, the knock-out of these genes suggests that SARS-CoV-2 originates from a distinct animal reservoir, explaining why several of its genes are not essential and may be knocked-out in humans, minks and pangolins. Similarly, ORF8 truncation has been hypothesized to have occurred for HCoV-229E in humans after zoonotic transmission from bats or intermediate hosts^28^. Functionally, ORF8 may help the virus to adapt to new hosts by facilitating immune evasion due to functional mimicry with immunological molecules^29^. This function may not remain critical as soon as the virus is well adapted to its new hosts. Interestingly, evolutionary changes in virulence observed for myxoma virus after its introduction as a biological control in rabbits of Australia were often associated with losses of gene functions^30^.

Regarding the spread of these Marseille-4 lineages, their average detection duration was approximately 4 months, indicating that the accumulation of mutations beyond 8 was associated with a vanishing. This accumulation of mutations is associated with genetic heterogeneity and increased diversity. This leads to a funnel-shaped evolution of the viral population. Overall, several beneficial mutations at unpredicted sites increase fitness, while the accumulation of other mutations results in decreased fitness, loss of clonality and vanishing of the lineage. We speculate that the observed heterogenicity of a lineage may predict the end of the outbreak.

## Data Availability

The dataset generated and analyzed during the current study are available in the GISAID database (https://www.gisaid.org/).

## Methods

### Patients

Patients included in the present study were those identified as infected with the SARS-CoV-2 Marseille-4 variant^1,2^. The present study has been approved by the ethics committee of University Hospital Institute (IHU) Méditerranée Infection (N°2020-016-3). Epidemiological and clinical data were retrieved for patients registered in the Assistance Publique-Hôpitaux de Marseille (APHM) information system. Access to the patients’ biological and registry data issued from this system was approved by the data protection committee of APHM and was recorded in the European General Data Protection Regulation registry under number RGPD/APHM 2019-73. Statistical processes were performed using R software version 4.0.2 (https://cran.r-project.org/). A p< 0.05 was considered statistically significant.

### SARS-CoV-2 genotyping

SARS-CoV-2 genotyping was performed from nasopharyngeal samples tested between 1 July 2020 and 30 April 2021 (10 months) at the IHU Méditerranée Infection Institute (https://www.mediterranee-infection.com/). Next-generation sequencing was performed when the cycle threshold value (Ct) of the qPCR used to diagnose SARS-CoV-2 infection was <30. When the Ct was ≥30 and in any case in the absence of an available genome sequence, the genotype was determined for SARS-CoV-2-positive specimens using variant-specific qPCR, as previously described^1,2,3^. Viral RNA was extracted from 200 µL of nasopharyngeal swab fluid using the EZ1 Virus Mini Kit v2.0 and the EZ1 Advanced XL instrument (Qiagen, Courtaboeuf, France) or the KingFisher Flex system (Thermo Fisher Scientific, Waltham, MA, USA) following the manufacturer’s instructions. SARS-CoV-2 genome sequences were obtained as follows: by next-generation sequencing using Illumina technology, the Nextera XT paired end strategy and the MiSeq instrument (Illumina Inc., San Diego, CA, USA) since February 2020, as previously described^2^; using the Illumina COVID-seq protocol and the NovaSeq 6000 instrument (Illumina Inc.) since April 2021; or using Oxford Nanopore technology (ONT) and the GridION instrument (Oxford Nanopore Technologies Ltd., Oxford, UK), as previously described^2^. Next-generation sequencing with ONT was performed without or with (since March 2021) synthesized cDNA amplification using a multiplex PCR protocol with ARTIC nCoV-2019 v3 Panel primers purchased from Integrated DNA technologies (IDT, Coralville, IA, USA) according to the ARTIC procedure (https://artic.network/), as previously described. Postextraction, viral RNA was reverse-transcribed using SuperScript IV (Thermo Fisher Scientific) before cDNA second strand synthesis with Klenow Fragment DNA polymerase (New England Biolabs, Beverly, MA, USA) when performing NGS using the Illumina MiSeq instrument (Illumina Inc.)^2^, LunaScript RT SuperMix kit (New England Biolabs) when performing NGS with the ONT, or according to the COVIDSeq protocol (Illumina Inc.) following the manufacturer’s recommendations. The generated cDNA was purified using Agencourt AMPure XP beads (Beckman Coulter, Villepinte, France) and quantified using a Qubit 2.0 fluorometer (Invitrogen, Carlsbad, CA, USA).

### Assembly and analyses of genome sequences

Genome sequences were assembled by mapping on the SARS-CoV-2 genome GenBank accession no. NC_045512.2 (Wuhan-Hu-1 isolate) using CLC Genomics workbench v.7 (with the following thresholds: 0.8 for coverage and 0.9 for similarity) (https://digitalinsights.qiagen.com/) as previously described^2^ or Minimap2 (https://github.com/lh3/minimap2)^4^. Samtools (https://www.htslib.org/) was used for soft clipping of Artic primers (https://artic.network/) and to remove sequence duplicates^5^. Consensus genomes were generated using CLC Genomics workbench v.7 and Sam2consensus (https://github.com/vbsreenu/Sam2Consensus) through a first in-house script written in Python language (https://www.python.org/). Mutation detection was performed using the Nextclade tool (https://clades.nextstrain.org/) and freebayes (https://github.com/freebayes/freebayes)^6^ using a mapping quality score of 20 and results filtered by the Python script based on major nucleotide frequencies ≥ 70% and nucleotide depths ≥ 10 (when sequence reads were generated on the NovaSeq Illumina instrument (Illumina Inc.)) or ≥ 5 (when sequence reads were generated on the MiSeq Illumina instrument). SARS-CoV-2 genotyping was performed using a second in-house script written in Python language (https://www.python.org/) by comparing mutation patterns with those of our database of SARS-CoV-2 variants. Nextstrain clades and Pangolin lineages provided in the present study were determined using the Nextclade web application (https://clades.nextstrain.org/)^7,8^ and Pangolin web application (https://cov-lineages.org/pangolin.html)^9^, respectively. The sequences described in the present study have been deposited in the GISAID sequence database (https://www.gisaid.org/)^10^ and can be retrieved online using the GISAID online search tool with “Marseille” as a keyword, and then selecting sequence names containing “IHU” or “MEPHI”. The sequences have also been deposited in the IHU Marseille Infection website: https://www.mediterranee-infection.com/tout-sur-le-coronavirus/sequencage-genomique-sars-cov-2/.

### Phylogenetic reconstruction and definition and naming of Marseille-4 subvariants and lineages

Phylogenetic reconstruction based on SARS-CoV-2 genomes were performed for genome sequences >24,000 nucleotides using the 3extstrain/ncov tool (https://github.com/nextstrain/ncov) and then were visualized using Auspice (https://docs.nextstrain.org/projects/auspice/en/stable/).

### Structural analysis of the untruncated and truncated ORF8 protein

A structural model of the ORF8 protein was generated from pdb file 7JTL^11^. The gaps in the crystal structure were fixed by incorporating the missing amino acids with the Robetta protein structure prediction tool^12^, followed by energy minimization with the Polak-Ribière algorithm as previously reported^13^. Mutant and truncated proteins were then generated with Swiss-PdbViewer^14^ and submitted to several rounds of energy minimization as described^15^.

### Evolution of Marseille-4 subvariants and lineages and time dynamics of SARS-CoV-2 mutation accumulation

Duration of circulation of the different subvariants and lineages was calculated using the differences between the 5^th^ and 95^th^ percentiles of sampling dates; this allowed considering the time periods during which the subvariants and lineages had a significant incidence. Time dynamics of mutation accumulation were analysed using locally estimated scatterplot smoothing (loess) for regression fitting. Latent structural changes in mutation distributions were retrieved using change point analysis of mean and variance with binary segmentation method and Schwartz information criterion penalty associated with a penalty threshold of 0.05^16^. We assessed the epidemiological capabilities of subvariants using the early stages of each epidemic curve. During this exponential phase, the size of the susceptible population may be considered as constant, and the cumulated number of cases exponentially increases at an approximately constant rate that was defined as the growth rate. After logarithmic transformation, the cumulated number of cases followed a linear model as follows: ln(It) = ln(I0) + Λt, where It is the cumulated incidence at time t, I0 is the initial number of cases, and Λ is the regression slope and the growth rate. From Λ, the reproduction rate, R, may be easily retrieved using the following equation^17^: R = (I+ΛD)(1+ΛD’), where D and D’ are the average infectious and pre-infectious periods (according to the SEIR model). Here, we set D at 9.3^18^ and d’ at 3.3^19^. To calculate the growth rate, we used Chow’s F test to determine the inflexion point of the logarithm of cumulated number of cases, which corresponded to the end of the exponential phase. Then, we applied a linear model for this phase to obtain the regression slope (i.e. the growth rate) and its 95% confidence interval. Statistical processes were performed using R software version 4.0.2 (https://cran.r-project.org/). All statistical conclusions were made using a 0.05 threshold.

## Acknowledgments

This work was supported by the French Government under the “Investments for the Future” program managed by the National Agency for Research (ANR), Méditerranée-Infection 10-IAHU-03 and the Emergen Consortium (https://www.santepubliquefrance.fr/dossiers/coronavirus-covid-19/consortium-emergen). We also acknowledge support from the Région Provence Alpes Côte d’Azur and European funding FEDER PRIMMI (Fonds Européen de Développement Régional-Plateformes de Recherche et d’Innovation Mutualisées Méditerranée Infection), FEDER PA 0000320 PRIMMI.

We are thankful to Mamadou Beye, Emilie Burel, Elsa Prudent, Céline Gazin, Sofiane Bakour, Laurence Thomas, Séverine Guillon, Véronique Filosa, Ludivine Bréchard, and Claudia Andrieu for their technical help.

## Author contributions

Study conception and design: D.R., P.C., B.L.S., P.E.F, P.G.. Materials, data and analysis tools: P.C., P.G., J.D., H.C., M.B., L.D., A.L., J.C.L., M.M., N.Y., J.F., P.E.F.. Data analyses: D.R., P.C., H.C., J.D., M.B., L.D., N.Y., J.F., B.L.S., P.E.F.. Writing of the first draft of the manuscript: P.C. and D.R.. Critical reviews of and revisions to the manuscript: D.R., P.C., P.G., P.P., P.F., J.F., B.L.S., P.E.F.. All authors approved the final manuscript. Supervision: D.R..

## Competing interests

D.R. is a scientific board member of Eurofins company, a founder of a microbial culture company (Culture Top), and was a consultant for Hitachi High-Technologies Corporation, Tokyo, Japan from 2018 to 2020. All other authors do not have any competing interest to declare. Funding sources had no role in the design and conduct of the study; collection, management, analysis, and interpretation of the data; and preparation, review, or approval of the manuscript.

## Funding

French Government under the “Investments for the Future” program managed by the National Agency for Research (ANR), no. Méditerranée-Infection 10-IAHU-03, and the “Emergen Consortium” (https://www.santepubliquefrance.fr/dossiers/coronavirus-covid-19/consortium-emergen).; Région Provence Alpes Côte d’Azur and European funding FEDER PRIMMI (Fonds Européen de Développement Régional-Plateformes de Recherche et d’Innovation Mutualisées Méditerranée Infection), no. FEDER PA 0000320 PRIMMI.

## Extended data figure/table legends

See Supplementary Material.

## Supplementary material

### Supplementary figure legends

**Supplementary Figure S1a-f.**
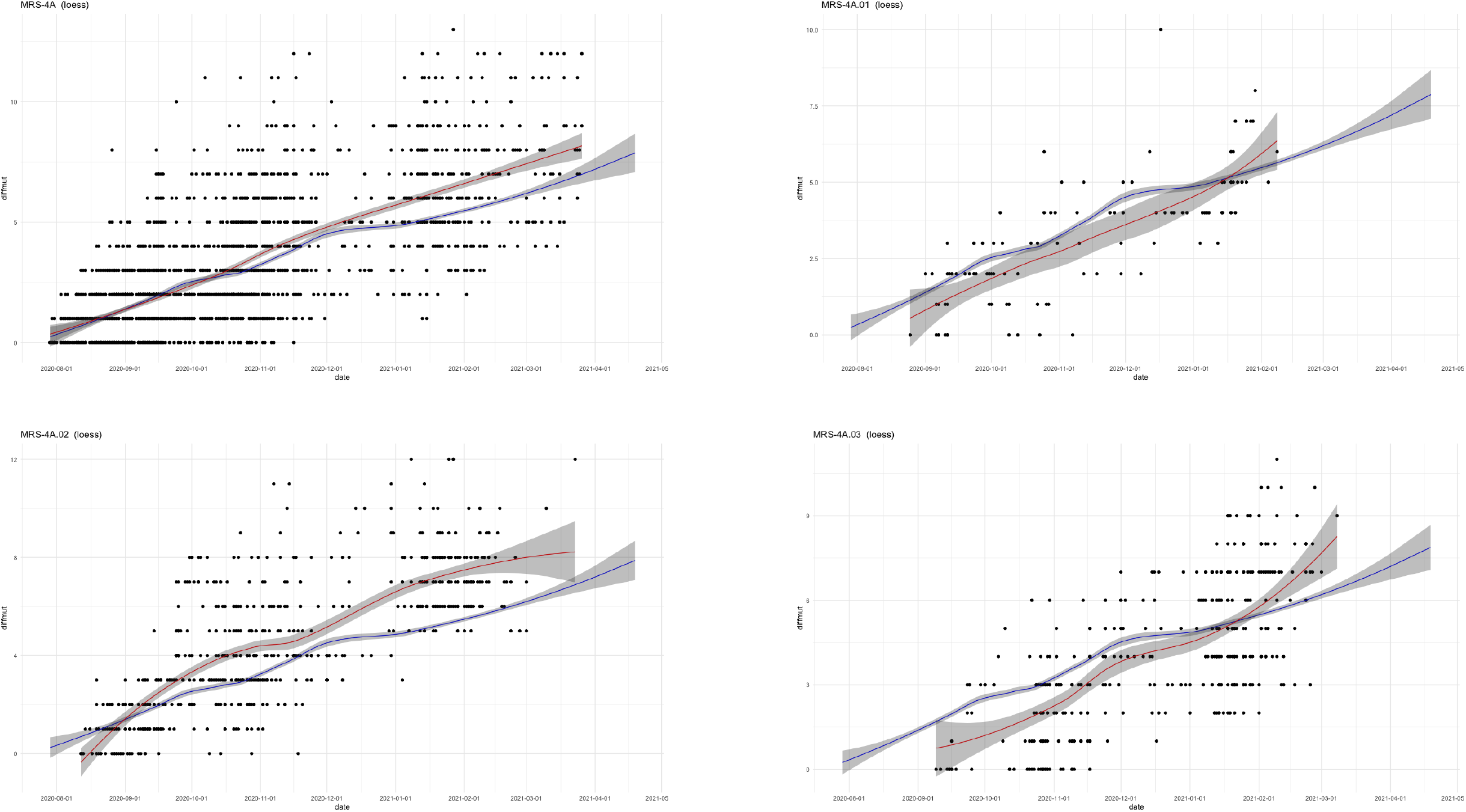
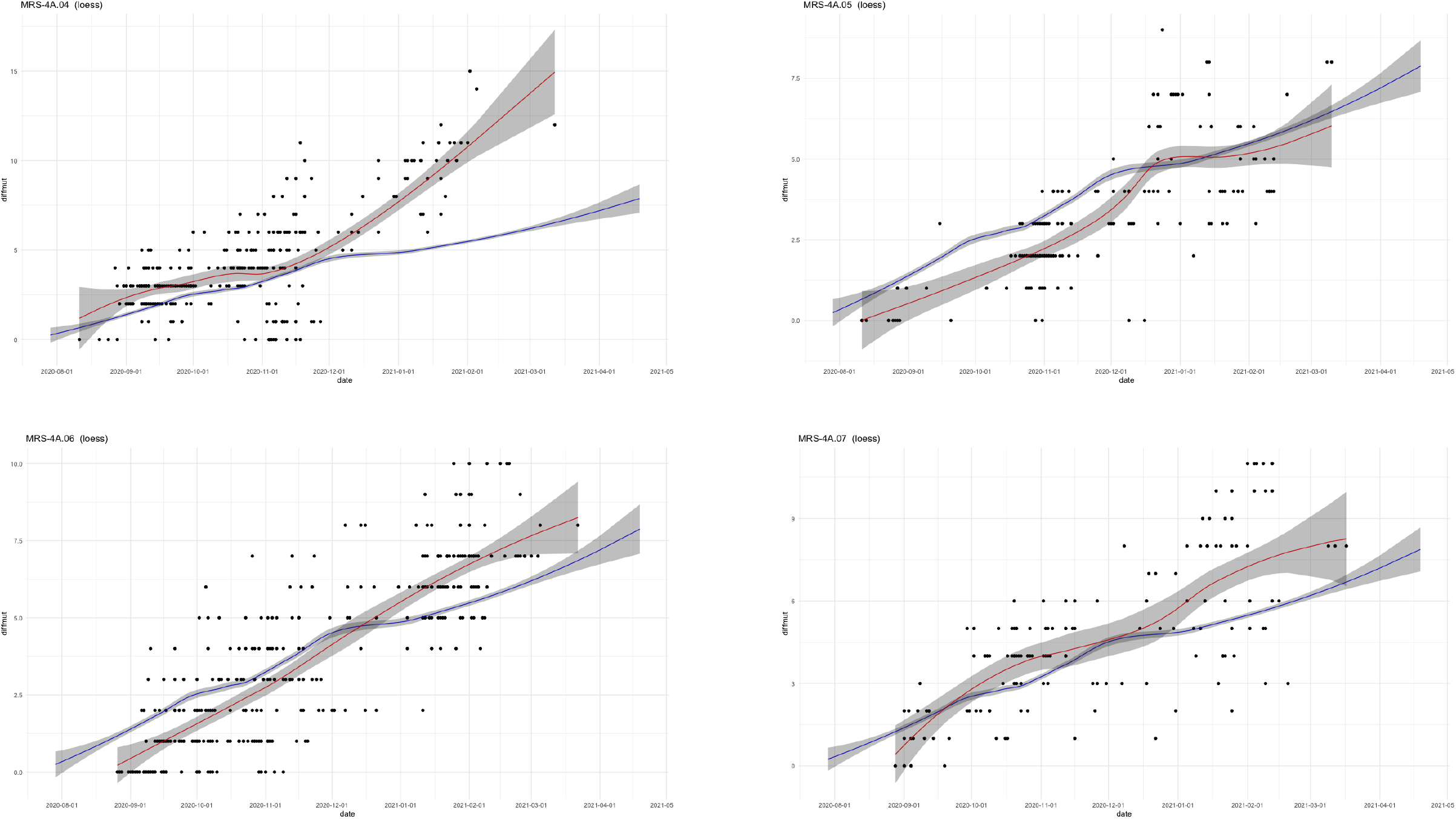
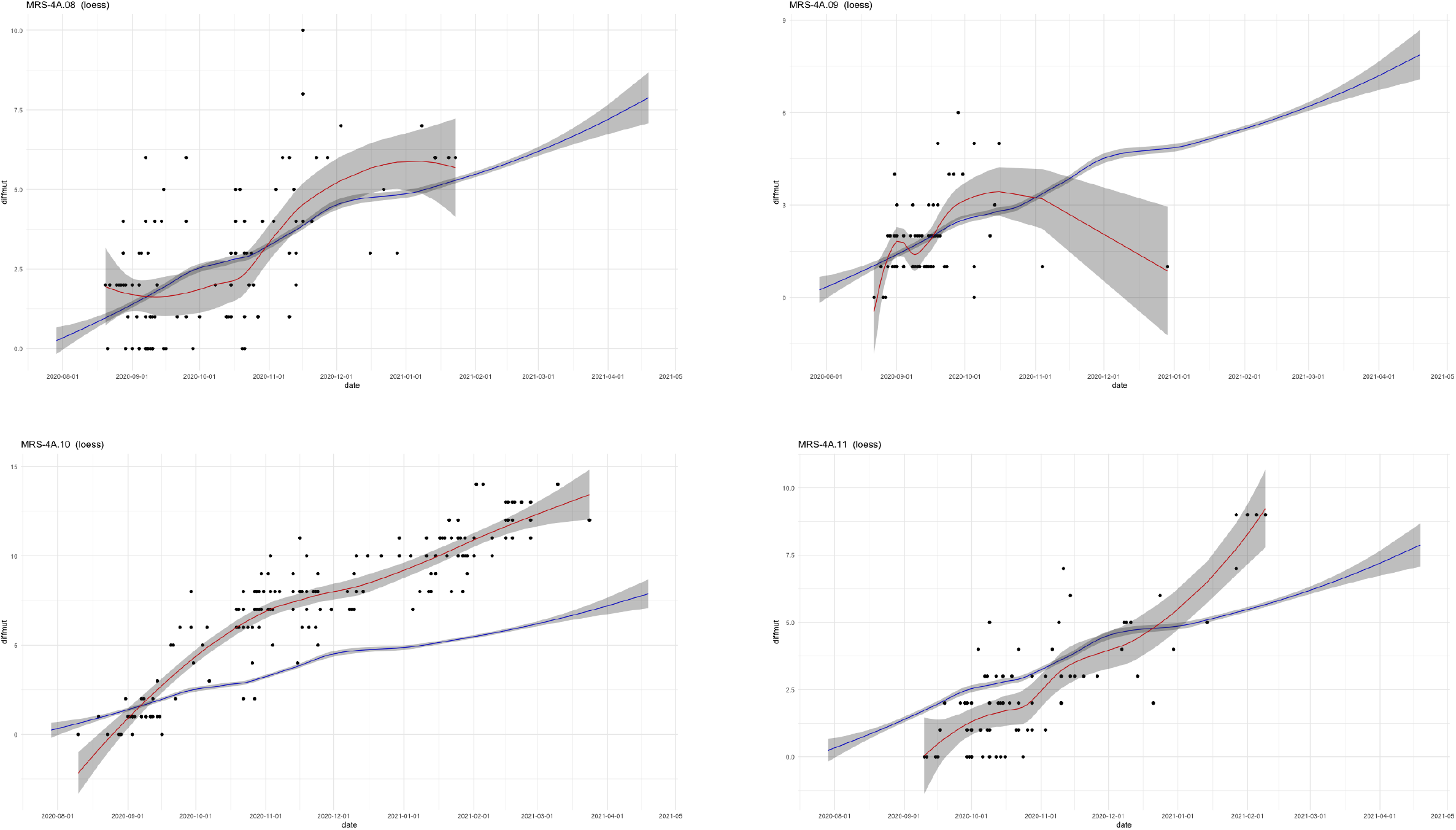
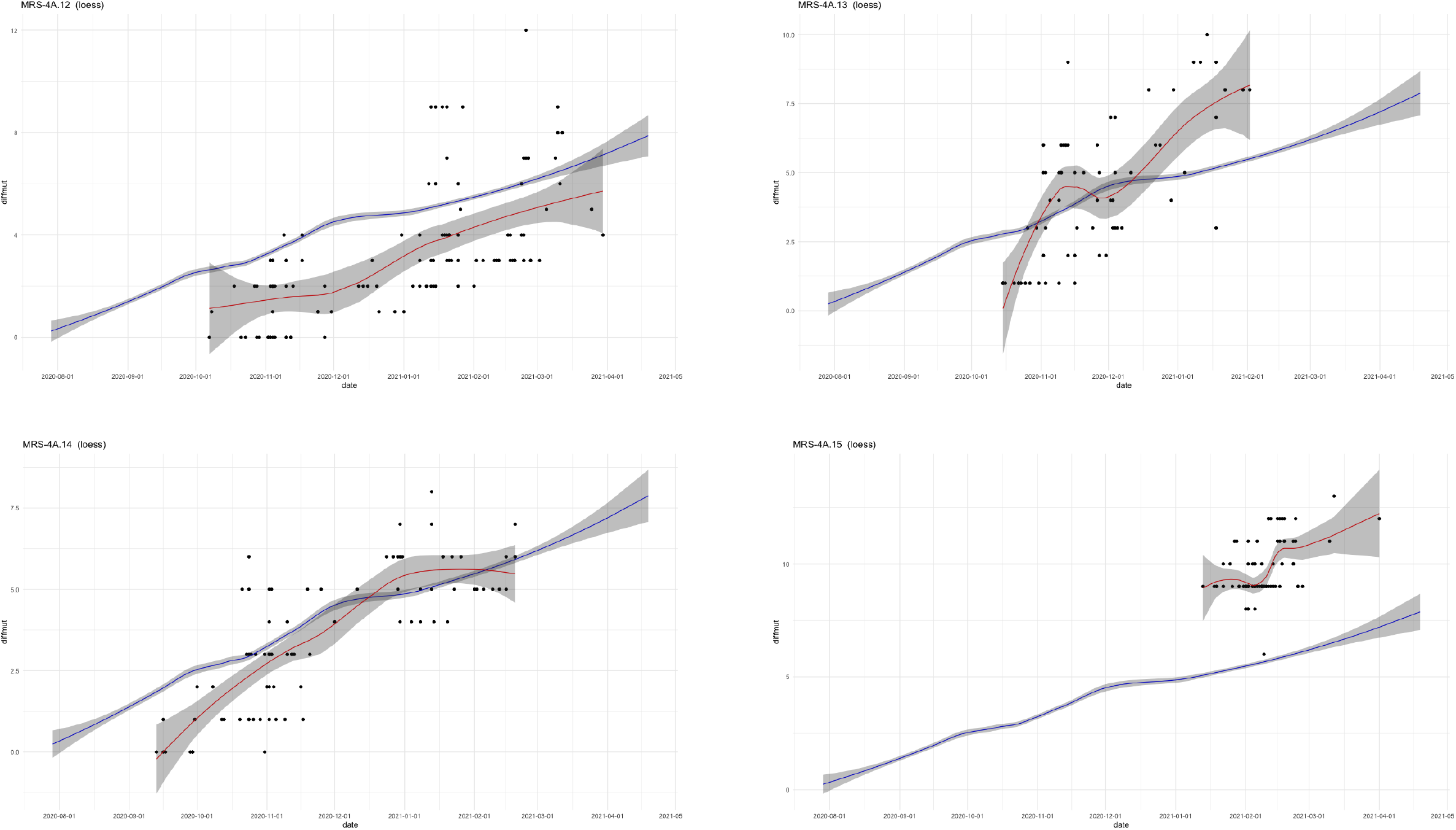
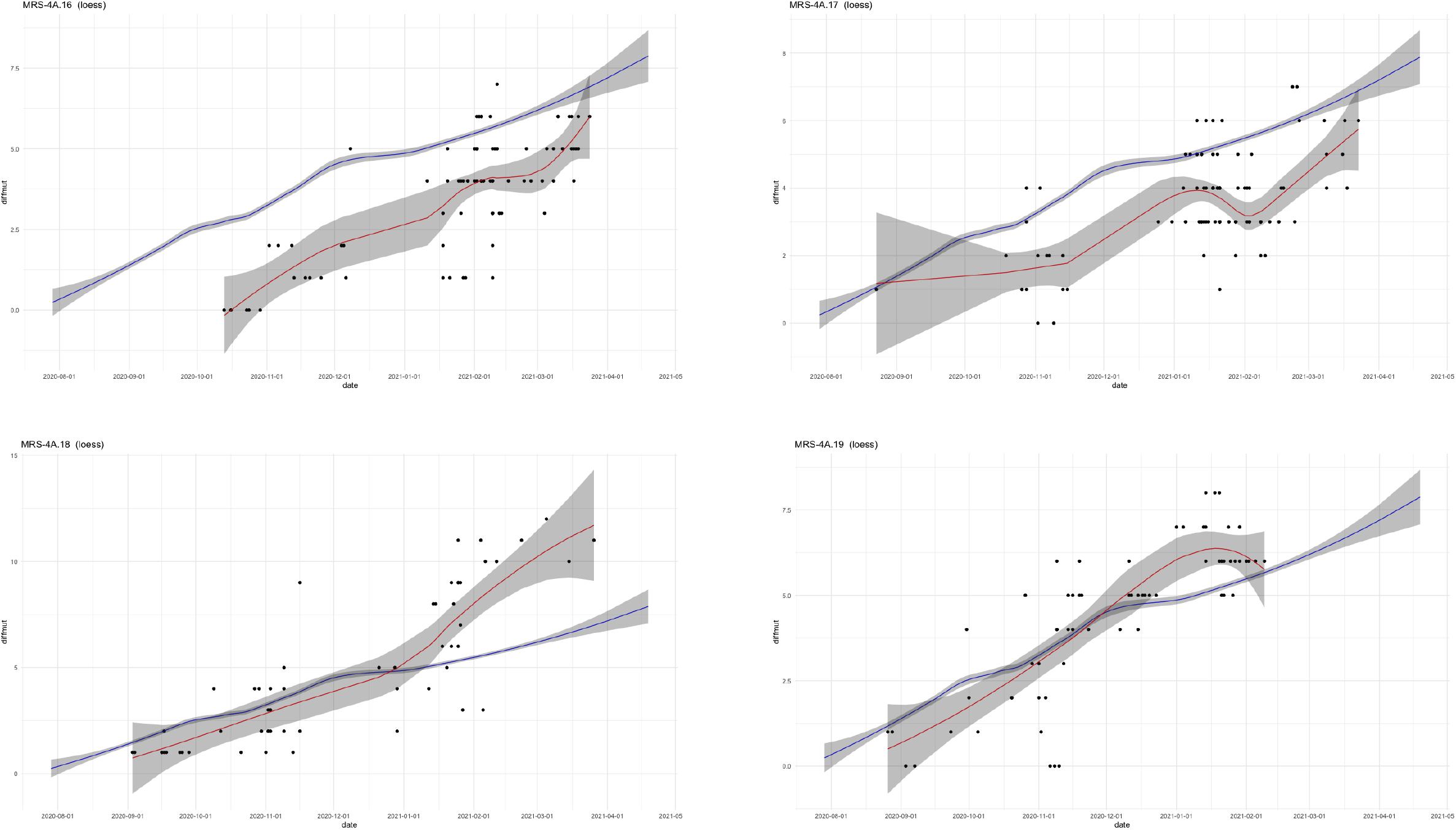
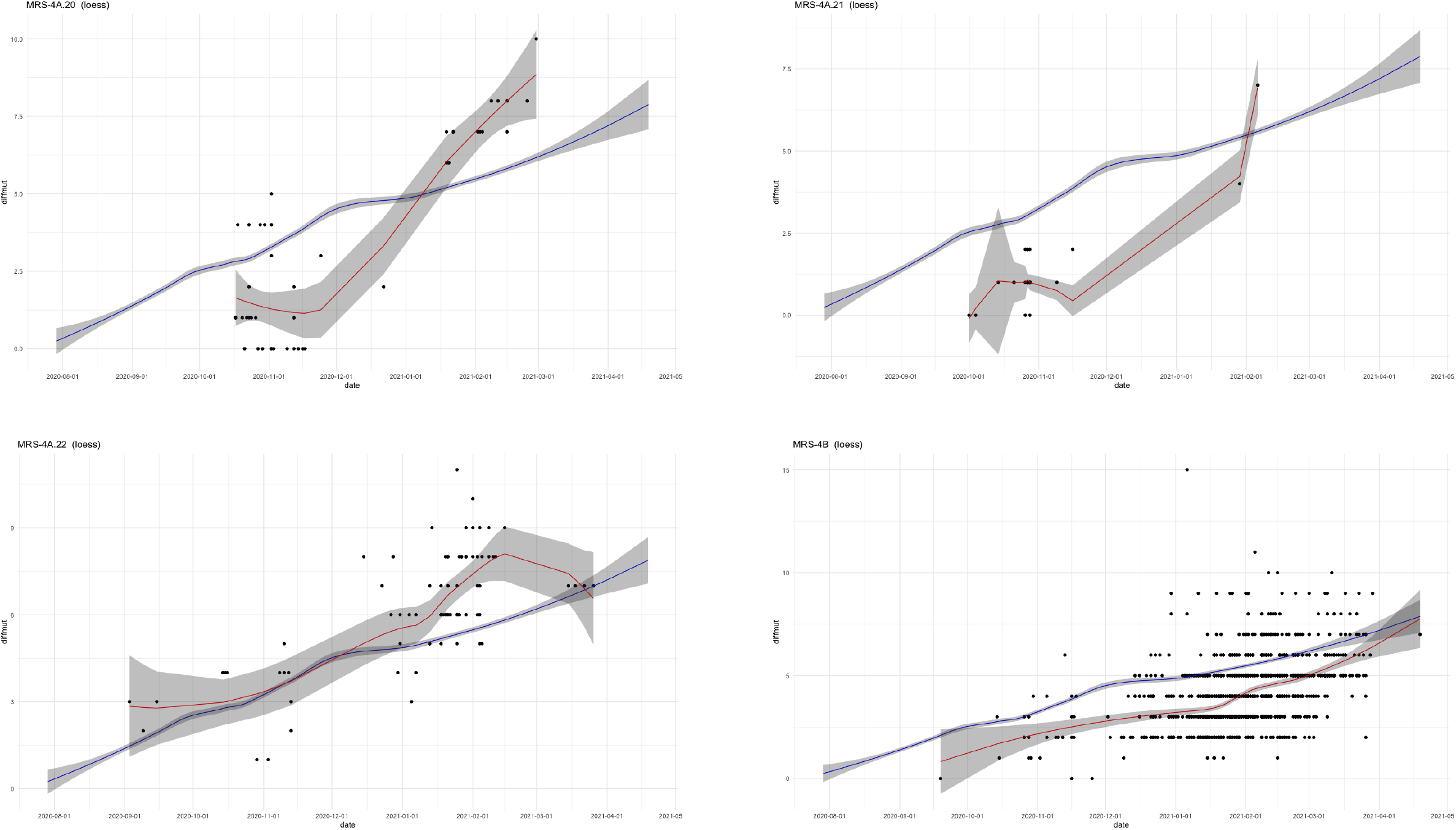
Time series for the different Marseille-4 subvariants and lineages of the number of mutations showing both the loess regression curve and the overall Marseille-4 loess regression curve, with their 95% confidence intervals. MRS, Marseille.

**Supplementary Figure S2.**
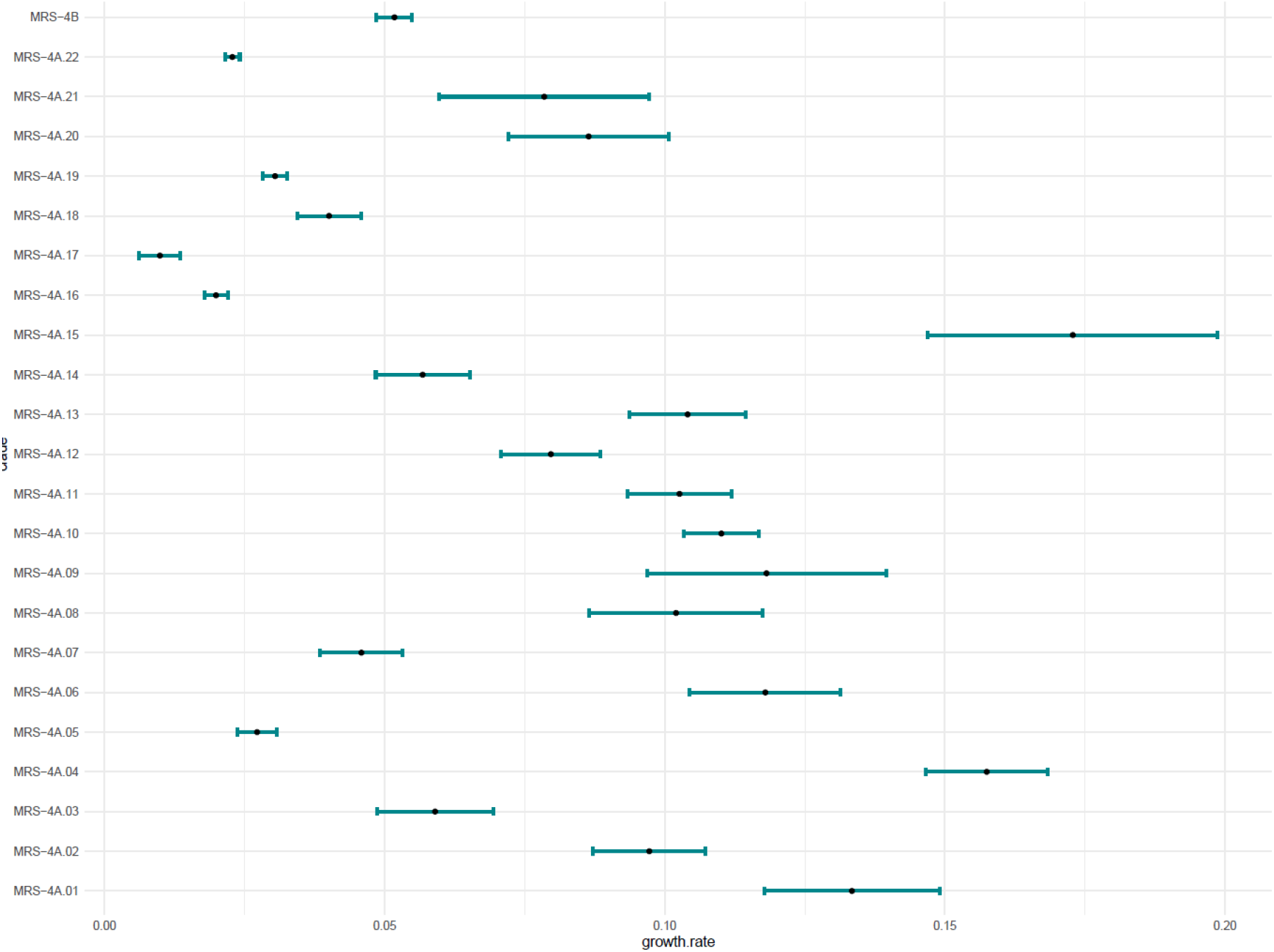
Forest plot like representation of the growth rates of subvariants and lineages with their 95% confidence intervals.

**Supplementary Figure S3.**
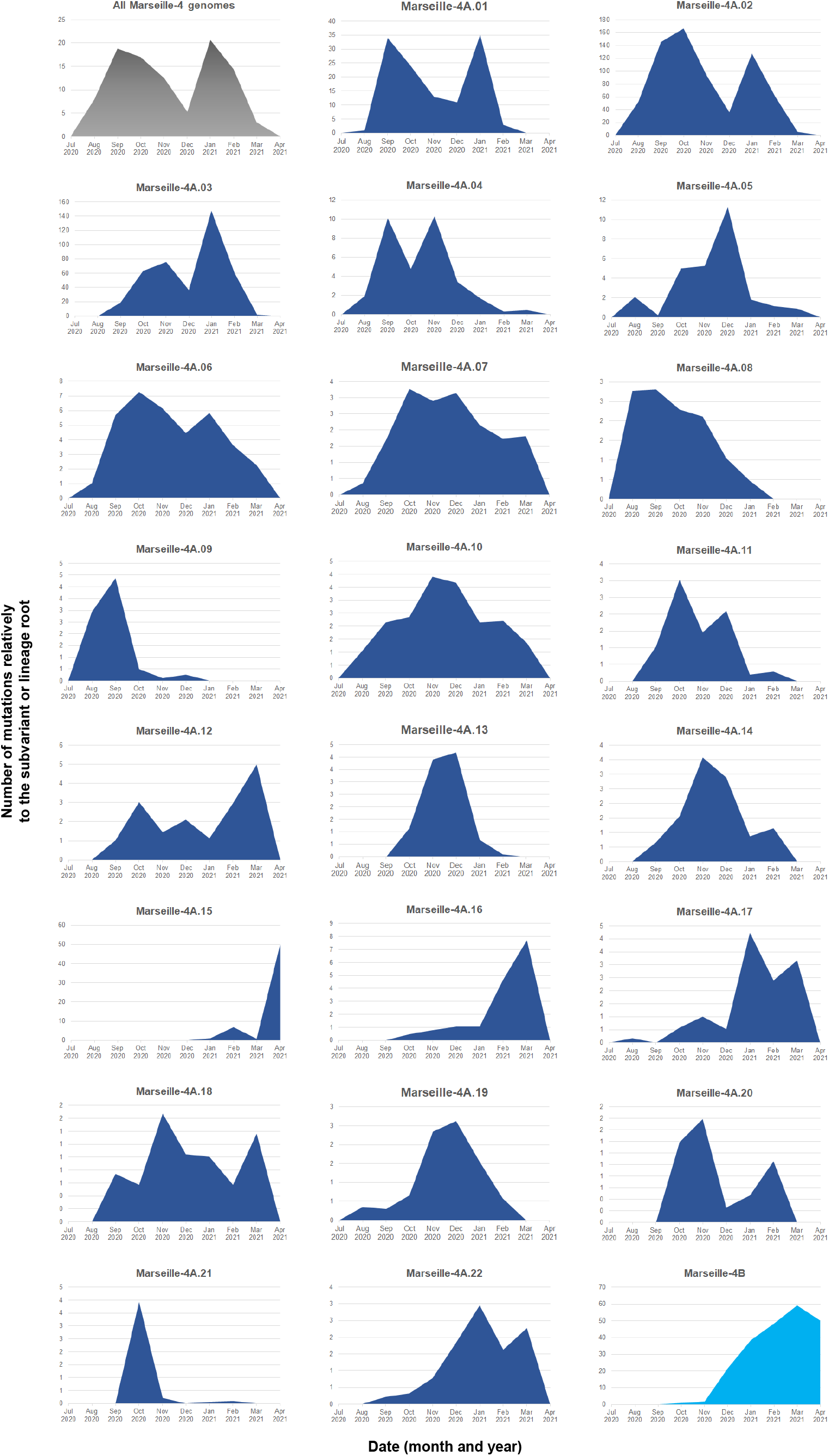
Temporal distribution of the genome sequences of SARS-CoV-2 Marseille-4 subvariants and lineages obtained from patients diagnosed with SARS-CoV-2 infection at IHU Méditerranée Infection.

### Supplementary Tables

**Supplementary Table S1.** List of nucleotide and amino acid changes associated with the onset and expansion of Marseille-4A and Marseille-4B subvariants and lineages.

| Marseille-4<br>subvariants and<br>lineages | Nucleotide changes (hallmark mutations) | Amino acid changes (hallmark mutations) |
| --- | --- | --- |
| Marseille-4A.01 | C23191U | - |
| Marseille-4A.02 | G28086U | ORF8: A65S |
| Marseille-4A.03 | G487A | - |
| Marseille-4A.04 | C222U | - |
| Marseille-4A.05 | C3096U; C23188U | ORF1a: S944L; - |
| Marseille-4A.06 | G29701A | - |
| Marseille-4A.07 | C27434U | ORF7a: T14I |
| Marseille-4A.08 | G571A | - |
| Marseille-4A.09 | G2600U | ORF1a: V779F |
| Marseille-4A.10 | G27877U | ORF7b: C41F |
| Marseille-4A.11 | U26442C; G29511U | -; N: S413I |
| Marseille-4A.12 | C503U; G28178U | ORF1a: P80S; ORF8: L95F |
| Marseille-4A.13 | C1551U; A11822G; C13548U; C22388U; G27816A | ORF1a: A429V; ORF1a: I3853V; -; -; ORF7b: V21I |
| Marseille-4A.14 | G7925U | ORF1a: A2554S |
| Marseille-4A.15 | C9866A; C26060U | ORF1a: L3201I; ORF3a: T223I |
| Marseille-4A.16 | G25340A; C29445U | S: D1260N; N: T391I |
| Marseille-4A.17 | G28655U | N: D128Y |
| Marseille-4A.18 | G26062U | ORF3a: G224C |
| Marseille-4A.19 | C6555U | ORF1a: A2097V |
| Marseille-4A.20 | A4870G | - |
| Marseille-4A.21 | C3743U | - |
| Marseille-4A.22 | U5071C; G8102U | -; ORF1a: V2613F |
| Marseille-4B | G204U; C5622U; C9924U; G17488A; G28083U; G29778U | -; -; ORF1a: A3220V; ORF1b: E1341K; ORF8: E64*; - |

See also Figure 3d; -, no amino acid change; \*, Stop codon

**Supplementary Table S2.** Time dynamics of Marseille-4 subvariants and lineages.

| Subvariants and lineages | Number of genomes | Change point (weeks) | Growth rate | Lower limit of growth rate (95% confidence interval) | Upper limit of growth rate (95% confidence interval) | p-value * |
| --- | --- | --- | --- | --- | --- | --- |
| Marseille-4A.01 | 121 | 32 | 0.133 | 0.118 | 0.149 | 3.08E-17 |
| Marseille-4A.02 | 685 | 35 | 0.097 | 0.087 | 0.107 | 8.61E-20 |
| Marseille-4A.03 | 404 | 48 | 0.059 | 0.049 | 0.069 | 4.97E-15 |
| Marseille-4A.04 | 339 | 34 | 0.157 | 0.147 | 0.168 | 7.47E-25 |
| Marseille-4A.05 | 207 | 76 | 0.027 | 0.024 | 0.031 | 1.14E-24 |
| Marseille-4A.06 | 374 | 32 | 0.118 | 0.104 | 0.131 | 1.56E-17 |
| Marseille-4A.07 | 157 | 52 | 0.046 | 0.038 | 0.053 | 4.81E-17 |
| Marseille-4A.08 | 112 | 33 | 0.102 | 0.087 | 0.117 | 1.78E-14 |
| Marseille-4A.09 | 84 | 33 | 0.118 | 0.097 | 0.140 | 1.66E-12 |
| Marseille-4A.10 | 171 | 35 | 0.110 | 0.103 | 0.117 | 3.63E-27 |
| Marseille-4A.11 | 78 | 30 | 0.103 | 0.093 | 0.112 | 1.85E-19 |
| Marseille-4A.12 | 131 | 27 | 0.080 | 0.071 | 0.089 | 4.24E-16 |
| Marseille-4A.13 | 76 | 25 | 0.104 | 0.094 | 0.114 | 2.06E-16 |
| Marseille-4A.14 | 96 | 42 | 0.057 | 0.048 | 0.065 | 1.08E-16 |
| Marseille-4A.15 | 91 | 19 | 0.173 | 0.147 | 0.199 | 8.18E-11 |
| Marseille-4A.16 | 99 | 106 | 0.020 | 0.018 | 0.022 | 5.42E-35 |
| Marseille-4A.17 | 114 | 66 | 0.010 | 0.006 | 0.014 | 1.27E-06 |
| Marseille-4A.18 | 60 | 57 | 0.040 | 0.034 | 0.046 | 6.89E-20 |
| Marseille-4A.19 | 74 | 68 | 0.030 | 0.028 | 0.033 | 3.45E-38 |
| Marseille-4A.20 | 52 | 24 | 0.086 | 0.072 | 0.101 | 1.81E-11 |
| Marseille-4A.21 | 52 | 27 | 0.078 | 0.060 | 0.097 | 6.15E-09 |
| Marseille-4A.22 | 87 | 119 | 0.023 | 0.021 | 0.024 | 1.30E-62 |
| Marseille-4B | 1,319 | 94 | 0.052 | 0.049 | 0.055 | 5.02E-52 |

\* P-values are for the statistical significance of the growth rate

**Supplementary Table S3.** Number of genomes, number of mutations and time duration for Marseille-4 subvariants and lineages.

| Subvariants and lineages | Number of genomes | Total number of mutations |  |  |  | Number of additional mutations relatively to the root |  |  |  | Raw data |  |  | 5<sup>th</sup> and 95<sup>th</sup> percentiles |  |  |
| --- | --- | --- | --- | --- | --- | --- | --- | --- | --- | --- | --- | --- | --- | --- | --- |
|  |  | Mean | Standard deviation | Minimum | Maximum | Mean | Standard deviation | Minimum | Maximum | Minimum date | Maximum date | Duration (months) | Minimum date | Maximum date | Duration (months) |
| Marseille-4A | 2 470 | 21,9 | 3,0 | 14 | 35 | 3,0 | 2,6 | 0 | 13 | 29/07/2020 | 26/03/2021 | 7,9 | 14/08/2020 | 04/02/2021 | 5,7 |
| Marseille-4A.01 | 121 | 22,9 | 2,7 | 17 | 29 | 3,2 | 2,1 | 0 | 10 | 25/08/2020 | 08/02/2021 | 5,5 | 07/09/2020 | 25/01/2021 | 4,6 |
| Marseille-4A.02 | 685 | 24,6 | 3,3 | 15 | 33 | 4,5 | 2,8 | 0 | 12 | 12/08/2020 | 23/03/2021 | 7,3 | 27/08/2020 | 08/02/2021 | 5,4 |
| Marseille-4A.03 | 404 | 24,3 | 3,0 | 16 | 32 | 4,1 | 2,5 | 0 | 11 | 09/09/2020 | 08/03/2021 | 5,9 | 01/10/2020 | 11/02/2021 | 4,4 |
| Marseille-4A.04 | 339 | 24,7 | 2,9 | 19 | 36 | 4,0 | 2,7 | 0 | 15 | 11/08/2020 | 12/03/2021 | 7,0 | 02/09/2020 | 14/01/2021 | 4,4 |
| Marseille-4A.05 | 207 | 24,6 | 2,2 | 17 | 31 | 3,1 | 1,9 | 0 | 9 | 11/08/2020 | 10/03/2021 | 6,9 | 28/08/2020 | 04/02/2021 | 5,2 |
| Marseille-4A.06 | 374 | 23,9 | 3,2 | 14 | 31 | 3,7 | 2,7 | 0 | 10 | 26/08/2020 | 22/03/2021 | 6,8 | 08/09/2020 | 09/02/2021 | 5,1 |
| Marseille-4A.07 | 157 | 24,7 | 3,5 | 15 | 32 | 4,6 | 2,8 | 0 | 11 | 28/08/2020 | 17/03/2021 | 6,6 | 04/09/2020 | 12/02/2021 | 5,3 |
| Marseille-4A.08 | 112 | 23,0 | 2,5 | 17 | 30 | 2,7 | 2,2 | 0 | 10 | 20/08/2020 | 23/01/2021 | 5,1 | 26/08/2020 | 10/01/2021 | 4,5 |
| Marseille-4A.09 | 84 | 22,7 | 1,6 | 18 | 27 | 1,9 | 1,2 | 0 | 6 | 22/08/2020 | 29/12/2020 | 4,2 | 26/08/2020 | 10/10/2020 | 1,5 |
| Marseille-4A.10 | 171 | 27,3 | 4,2 | 18 | 35 | 7,3 | 3,7 | 0 | 14 | 10/08/2020 | 24/03/2021 | 7,4 | 03/09/2020 | 18/02/2021 | 5,5 |
| Marseille-4A.11 | 78 | 23,0 | 3,4 | 16 | 31 | 2,5 | 2,3 | 0 | 9 | 10/09/2020 | 09/02/2021 | 5,0 | 16/09/2020 | 27/01/2021 | 4,3 |
| Marseille-4A.12 | 131 | 24,7 | 2,6 | 19 | 34 | 3,2 | 2,4 | 0 | 12 | 07/10/2020 | 30/03/2021 | 5,7 | 16/09/2020 | 27/01/2021 | 4,3 |
| Marseille-4A.13 | 76 | 27,3 | 3,5 | 20 | 35 | 4,3 | 2,5 | 1 | 10 | 15/10/2020 | 02/02/2021 | 3,6 | 21/10/2020 | 19/01/2021 | 2,9 |
| Marseille-4A.14 | 96 | 23,4 | 2,7 | 15 | 29 | 3,5 | 2,0 | 0 | 8 | 13/09/2020 | 19/02/2021 | 5,2 | 28/09/2020 | 12/02/2021 | 4,5 |
| Marseille-4A.15 | 91 | 31,5 | 1,7 | 24 | 35 | 9,7 | 1,2 | 6 | 13 | 13/01/2021 | 01/04/2021 | 2,6 | 22/01/2021 | 24/02/2021 | 1,1 |
| Marseille-4A.16 | 99 | 25,2 | 2,2 | 18 | 29 | 3,6 | 1,7 | 0 | 7 | 13/10/2020 | 24/03/2021 | 5,3 | 28/10/2020 | 17/03/2021 | 4,6 |
| Marseille-4A.17 | 114 | 24,0 | 2,0 | 16 | 28 | 3,5 | 1,4 | 0 | 7 | 23/08/2020 | 23/03/2021 | 7,0 | 28/10/2020 | 08/03/2021 | 4,3 |
| Marseille-4A.18 | 60 | 25,0 | 3,9 | 18 | 33 | 4,8 | 3,5 | 1 | 12 | 03/09/2020 | 26/03/2021 | 6,7 | 16/09/2020 | 22/02/2021 | 5,2 |
| Marseille-4A.19 | 74 | 24,7 | 3,1 | 16 | 29 | 4,5 | 2,1 | 0 | 8 | 26/08/2020 | 09/02/2021 | 5,5 | 17/09/2020 | 01/02/2021 | 4,5 |
| Marseille-4A.20 | 52 | 23,4 | 4,1 | 16 | 31 | 3,4 | 3,1 | 0 | 10 | 17/10/2020 | 28/02/2021 | 4,4 | 17/10/2020 | 19/02/2021 | 4,1 |
| Marseille-4A.21 | 52 | 21,8 | 1,3 | 16 | 26 | 1,2 | 1,0 | 0 | 7 | 01/10/2020 | 06/02/2021 | 4,2 | 17/10/2020 | 12/11/2020 | 0,8 |
| Marseille-4A.22 | 87 | 27,7 | 2,5 | 17 | 33 | 6,2 | 1,9 | 1 | 11 | 03/09/2020 | 26/03/2021 | 6,7 | 15/10/2020 | 06/03/2021 | 4,7 |
| Marseille-4B | 1 319 | 27,4 | 2,1 | 18 | 36 | 4,1 | 1,6 | 0 | 15 | 29/07/2020 | 19/04/2021 | 8,7 | 22/12/2020 | 10/03/2021 | 2,6 |
| Total | 7 453 | 24,3 | 3,6 | 14 | 36 | 3,8 | 2,6 | 0 | 15 | 29/07/2020 | 19/04/2021 | 8,7 | 26/08/2020 | 23/02/2021 | 6,0 |

## Notes

### Funding Statement

French Government under the Investments for the Future program managed by the National Agency for Research (ANR), no. Mediterranee-Infection 10-IAHU-03, and the Emergen Consortium (https://www.santepubliquefrance.fr/dossiers/coronavirus-covid-19/consortium-emergen); Region Provence Alpes Cote d Azur and European funding FEDER PRIMMI (Fonds Europeen de Developpement Regional-Plateformes de Recherche et d'Innovation Mutualisees Mediterranee Infection), no. FEDER PA 0000320 PRIMMI.

### Author Declarations

The present study has been approved by the ethics committee of University Hospital Institute (IHU) Mediterranee Infection (No. 2020-016-3).

